# Symptom risk and critical event prediction models for orthostatic hypotension in a-Synucleinopathies: Utilizing dynamic blood pressure and cerebral blood flow velocity

**DOI:** 10.1101/2025.09.15.25335826

**Authors:** Yihong Gu, Hongxiu Chen, Fubo Zhou, Songwei Chen, Yujun Chen, Yingqi Xing

## Abstract

**Background:** Current clinical management for orthostatic hypotension (OH) in α-synucleinopathies is passive and lacks specificity, and objective tools for accurately assessing the true risk level in OH patients are lacking. Therefore, this study aimed to develop an objective tool to quantify OH severity, enabling individualized and precision-based clinical management of OH.

**Methods:** From January 2021 to March 2025, patients with OH at Beijing Xuanwu Hospital were assigned an OH Symptom-Based Risk Score (0–2 points) based on symptom severity. Active standing tests were performed to continuously collect data including blood pressure (BP) and cerebral blood flow velocity (CBFv). Two predictive models were developed: one to identify patients with versus without OH symptoms, and another to predict high-risk patients prone to critical events among symptomatic patients (scoring 1–2 points).

**Results:** Among 172 patients with OH (86 asymptomatic, 50%), symptomatic patients showed greater orthostatic reductions in BP and CBFv (both maximum and mean decreases, all P < 0.05) than asymptomatic patients. OH Symptom-Based Risk Scores were significantly correlated with all orthostatic BP and CBFv decline parameters (all P < 0.001). Binary logistic regression models successfully identified symptomatic patients and predicted patients at high risk for critical events among symptomatic patients, with bootstrap-validated accuracy exceeding 70% in multiple models. Orthostatic BP and CBFv decline parameters were independent predictors of OH.

**Conclusion:** The established models provide an objective tool for quantifying OH risk severity, showing significant potential for early clinical warning and optimization of OH management strategies.

## Introduction

Parkinson’s disease (PD) and multiple system atrophy (MSA) represent common neurodegenerative α-synucleinopathies^1^, and recent clinical management studies have emphasized non-motor symptoms of these diseases. Orthostatic hypotension (OH), a cardinal manifestation of chronic autonomic dysfunction, affects 19–43% of patients with PD and > 50% of patients with MSA^2,3^, imposing a growing burden on patients and society^4,5^. OH manifests as a drop in blood pressure (BP) during postural changes and is frequently accompanied by reduced cerebral perfusion. This condition increases risk of myocardial ischemia^3^, dementia^6^, with a 10-year mortality rate as high as 64%^7^. The severity of symptoms in patients with OH varies, with dizziness or blurred vision impairing daily function, while severe cases trigger critical events, including falls, syncope, or unconsciousness^3,8^. These events in turn increase fracture risk and reduce life expectancy in the older population. Notably, 50% patients with OH lack early symptoms and remain undiagnosed^9^. Moreover, approximately 40% individuals with severe BP decline show no symptoms yet sustain a multi-organ damage risk^3^. Current management also has critical limitations; therapy initiation relies on the presence of symptoms and neglects high-risk asymptomatic subgroups. Additionally, pharmacological intervention is typically delayed until nonpharmacological methods fail^3,10,11^, potentially enabling preventable catastrophic events. Consequently, the precise stratification of OH risk severity is essential for individualized treatment.

When patients with OH transition from the supine position to the standing position, blood redistributes to areas such as the lower limbs. This may lead to cerebral hypoperfusion in case of arterial baroreflex dysfunction, thereby inducing core OH symptoms such as dizziness, syncope, and falls. The human body possesses a dynamic cerebral autoregulation (dCA) mechanism to maintain stable cerebral blood flow^12,13^, which functions when the mean arterial pressure is within the range 60–150 mmHg^14^. However, in patients with OH, a substantial drop in BP may impair the dCA. Moreover, several studies have demonstrated an association between the magnitude of BP drop and OH symptoms. Justin et al.^15^observed cognitive decline in such patients, while Juraschek et al.^16^ found that the incidence of symptoms, such as dizziness and blurred vision, was positively correlated with the magnitude of BP decrease. A recent meta-analysis^17^ also indicated that symptomatic patients with OH exhibit a greater reduction in cerebral blood flow velocity (CBFv) compared to asymptomatic patients with OH, and the underlying mechanism may be associated with impaired dCA. Notably, previous studies^7,8^ have primarily focused on classic OH occurring within 3 min, often employing intermittent blood pressure measurements at 1-min intervals. However, according to the 2021 US and European consensus on OH^18^, it can be classified based on the time of onset as initial OH (within 15 s), classic OH (15 s–3 min), or delayed OH (> 3 min), with different subtypes exhibiting prognostic differences^3,19^. Gibbons et al.^7^ found that the 10-year mortality rate of classic OH was much higher than that of delayed OH, whereas Stephen et al.^16^ discovered that initial OH significantly increased the risk of short- and long-term OH symptoms compared to other OH subtypes. Moreover, OH can be classified as neurogenic (nOH) or non-nOH. nOH, which results from impaired baroreflex pathways leading to vasoconstrictor dysfunction, has a more severe long-term prognosis than non-nOH^8^. Furthermore, compared with patients with PD, patients with MSA have a higher incidence of OH and more severe functional impairment^1^. However, stratified studies focusing on risk events, such as falls and syncope, in symptomatic patients with OH are currently lacking. Consequently, it is necessary to analyze the independent predictors of dangerous OH symptoms to provide early warning.

Therefore, this study aimed to establish an objective tool for quantifying true OH risk severity, by identifying indicators associated with OH symptom severity and developing predictive models: one to distinguish symptomatic from asymptomatic patients, and another to identify high-risk patients susceptible to critical events among symptomatic cohorts. The results of this study will enable individualized and precision-based clinical management of OH.

## Methods

The current study was approved by the Ethics Committee of Beijing Xuanwu Hospital (Approval No. KS2025247). Informed consent was obtained from each patient before enrolment.

### Study population

This study included patients with clinically established or probable PD or MSA^20,21^ accompanied by OH, recruited at the Beijing Xuanwu Hospital, Capital Medical University, between January 2021 and March 2025. Comprehensive baseline clinical data were collected, including demographic characteristics, and medical history. Additionally, standardized clinical assessments, including the Movement Disorder Society-Unified Parkinson’s Disease Rating Scale (MDS-UPDRS), 39-item Parkinson’s Disease Questionnaire (PDQ-39), Mini-Mental State Examination (MMSE), and Montreal Cognitive Assessment (MoCA) were performed. The exclusion criteria were: 1) age under 18 years; 2) incomplete clinical documentation, 3) inadequate bilateral temporal window acoustic penetration, 4) comorbid conditions, such as cerebral infarction, atrial fibrillation, severe steno-occlusive disease of head or neck vasculature, or other confirmed dCA-impairing disorders, affecting dCA^22^; and 5) inability to complete active standing tests owing to postural transition limitations, communication barriers, or related mobility constraints.

### Active standing test

All participants abstained from alcohol, caffeine, nicotine, dopamine agents, and vasoactive medications for 24 h prior to assessment. Examinations were conducted during consistent morning hours in a temperature-controlled, silent room (20–24°C). After 10 min of rest in the supine position, the participants performed an active standing test (AST) comprising 10 min rest in the supine position, followed by 10 min of standing (Figure 1). Based on the 2021 US and European consensus on OH^18^, we categorized OH into three chronological subtypes: (1) initial OH, transient systolic blood pressure (SBP) decrease ≥ 40 mmHg or diastolic blood pressure (DBP) decrease ≥ 20 mmHg within 15 s of standing; (2) classic OH, SBP decrease ≥ 20 mmHg or DBP decrease ≥ 10 mmHg within 3 min of standing; (3) delayed OH, SBP decrease ≥ 20 mmHg or DBP decrease ≥ 10 mmHg occurring > 3 min after standing. The etiological classification was used to distinguish nOH from non-nOH. nOH was identified by a Δheart rate/ΔSBP ratio < 0.492 within 3 min after standing^23^.

**Figure 1.**
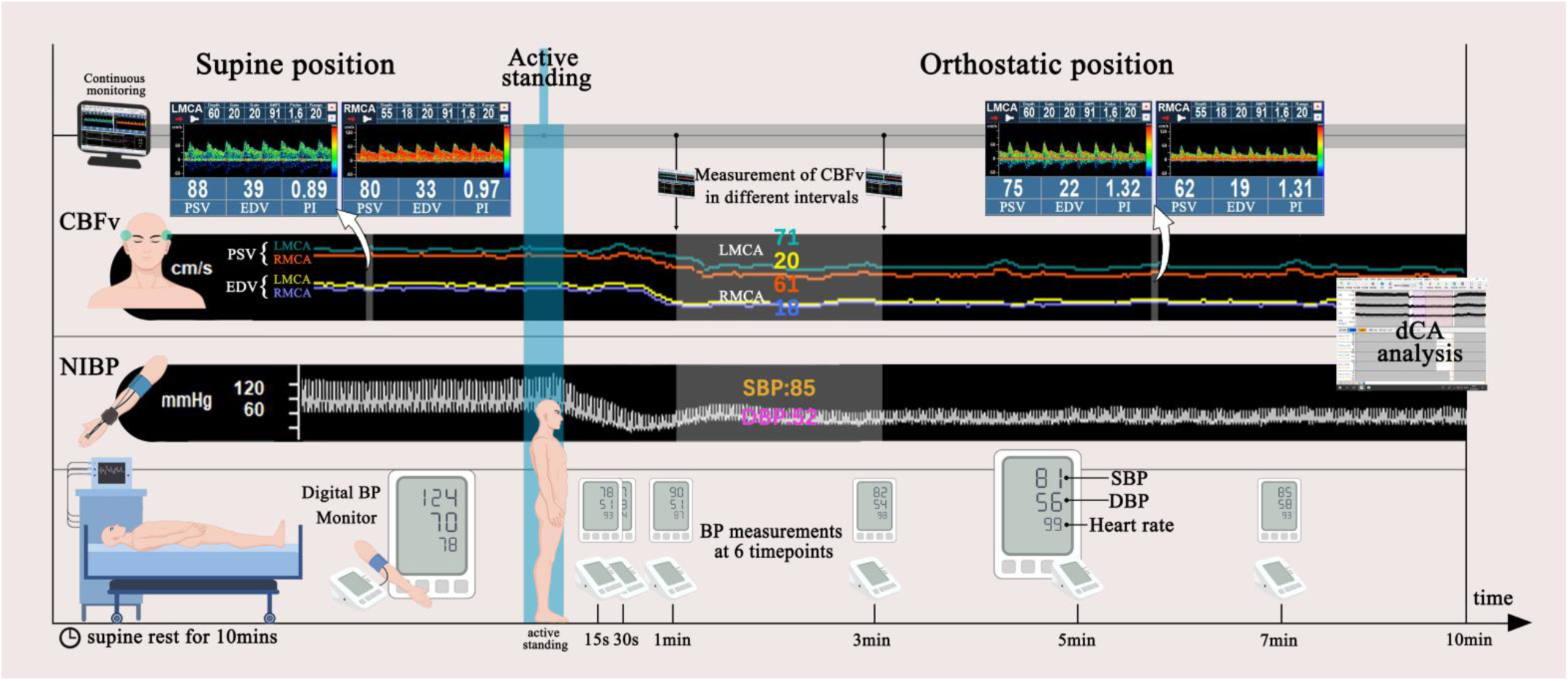
Illustration of the Active Standing Test Protocol. The protocol timeline (left to right) outlines the experimental flow and instrumental setup. Standard equipment included: computerized analysis software, 2-MHz transcranial Doppler probes for bilateral middle cerebral artery (MCA) cerebral blood flow velocity (CBFv) monitoring, continuous non-invasive beat-to-beat blood pressure (NIBP) recording, an automated oscillometric cuff for intermittent brachial blood pressure (BP) measurement (positioned on the contralateral upper limb), and continuous electrocardiographic heart rate monitoring. The protocol commenced with a 10-minute supine rest period, during which baseline hemodynamic parameters were acquired. Participants then actively stood and maintained an upright posture for 10 minutes. Average peak systolic velocity (PSV) and end-diastolic velocity (EDV) were computed over sequential post-standing intervals (0–15 s, 15–30 s, 30–60 s, 1–3 min, 3–5 min, 5–10 min). Systolic and diastolic BP (SBP/DBP) were recorded at fixed timepoints (15 s, 30 s, 1 min, 3 min, 5 min, and 7 min). Dynamic cerebral autoregulation (dCA) parameters were generated using transfer function analysis (TFA).

Baseline BP was measured in the brachial artery (Omron HBP1300; Kyoto, Japan) with the patient in the supine position three times. We strictly adhered to the 2022 TFA White Paper^24^ using an EMS-9D PRO (Delica Medical, Shenzhen, China) for dCA assessment to simultaneously record the NIBP and CBFv during the entire procedure. Continuous NIBP measurements were acquired using a servo-controlled plethysmograph positioned on the middle finger. Additionally, bilateral 2-MHz transcranial Doppler probes were fixed over the temporal windows using an adjustable head frame. CBFv was continuously measured in the bilateral middle cerebral arteries at depths of 50–60 mm. When the bilateral temporal windows permitted adequate measurements, mean CBFv was calculated as the average of the left and right measurements. No significant interhemispheric differences were observed in CBFv or dCA parameters (all P > 0.05). The recorded NIBP (input signals) and CBFv (output signals) data were used to calculate the dCA parameters based on TFA^25^, including the phase, absolute gain (cm/s/mmHg), normalized gain (%/mmHg), coherence at very low frequency (VLF, 0.02–0.07 Hz), low frequency (LF, 0.07–0.2 Hz), and high frequency (HF, 0.2–0.5 Hz). This phase represents the time delay in CBFV response to NIBP. The gain represents the moderation of BP oscillation magnitude by dCA, absolute gain captures actual magnitude changes in NIBP and CBFV, and normalized gain expresses their relative variance, irrespective of baseline levels. All analyses assumed system linearity, with a coherence approaching 1, indicating optimal linearity. The standing-position dCA data were exclusively analyzed, excluding epochs with coherence < 0.5. Only the VLF and LF band phases and gain results were reported because they are considered the most valuable evaluation metrics among the dCA parameters^24^. Impaired dCA manifests as decreased phase and increased gain values^24^.

### Orthostatic hemodynamic parameters

The orthostatic transition from the supine to the standing position induces real-time fluctuations in BP and cerebral perfusion. Therefore, SBP and DBP were measured at six post-standing intervals at 15 and 30 s, and 1, 3, 5, and 7 min to comprehensively characterize these dynamics. Subsequently, the CBFv parameters, peak systolic velocity (PSV), and end diastolic velocity (EDV) were averaged across six sequential intervals: 0–15 s, 15–30 s, 30– 60 s, 1–3 min, 3–5 min, and 5–10 min. Hemodynamic changes were quantified using the following two metrics: 1) maximum decreases (ΔSBPmax, ΔDBPmax, ΔPSVmax, ΔEDVmax) representing the largest observed reductions during measurements; and 2) mean decreases (ΔSBPmean, ΔDBPmean, ΔPSVmean, ΔEDVmean) reflecting average reductions across timepoints/intervals. This multitemporal monitoring protocol captured the evolving hemodynamic profile during orthostatic challenge, mirroring the mechanistic precision established in the background section.

### Symptom-based risk stratification of OH

Finally, a brief questionnaire adapted from a validated OH symptom questionnaire^26^ was administered to assess the presence of OH-related symptoms in the preceding month. Symptoms were rated on a 0–2 risk severity scale: 0 points (asymptomatic), 1 point (symptoms affecting daily living: dizziness, blurred vision, coat-hanger pain, and blacking out), and 2 points (symptoms associated with hazardous events: syncope, loss of consciousness, falls due to dizziness, and inability to walk due to dizziness). This OH Symptom-Based Risk Score was used as a reference to analyze its potential association with various parameters, including clinical demographics (e.g., age and sex), clinical scale scores, OH subtypes, type of synucleinopathy (PD or MSA), and hemodynamic parameters (BP and CBFv metrics). Subsequently, two predictive models were developed: one to discriminate between asymptomatic and symptomatic patients and the other to identify high-risk individuals prone to hazardous events within the symptomatic cohort.

### Statistical analysis

All data were analyzed using SPSS (version 27.0.1) and Python (version 3.11). Normality was first assessed using the Shapiro–Wilk test. Normally distributed continuous variables (mean ± SD) were analyzed by independent t-test or analysis of variance (ANOVA) with post-hoc tests. Non-normally distributed variables (median[IQR]) were analyzed using Mann–Whitney U, Kruskal–Wallis, or Wilcoxon tests as appropriate. Correlations were calculated using Pearson’s (normal) or Spearman’s (non-normal) coefficients. Categorical variables were compared using χ² or Fisher’s exact tests. Risk factors showing statistically significant differences (P < 0.05) in the univariate analyses (comparing relevant groups) for the two analytical goals, (1) identifying asymptomatic and symptomatic OH patients and (2) stratifying symptomatic patients by critical symptom, were subsequently analyzed for collinearity. Factors exhibiting a variance inflation factor > 10 were excluded from simultaneous inclusion in the multivariate model. Binary logistic regression analysis was conducted separately for each goal. Receiver operating characteristic (ROC) curves were constructed and the area under the ROC curve (AUC) was calculated. Finally, the predictive performance of the models was validated using a bootstrap resampling method (B = 1000). Figures were generated using Python (Matplotlib/Seaborn) and GraphPad Prism software. Statistical significance for all analyses was set at P < 0.05 (two-tailed).

## Results

### Comparison of baseline and OH-related parameters between asymptomatic and symptomatic patients

A total of 217 patients were enrolled in this study. After excluding 37 patients due to bilateral temporal window penetration failure, two due to acute cerebral infarction, four due to atrial fibrillation, and two due to internal carotid artery occlusion, a total of 172 patients were finally analyzed. The patients were divided into two groups based on the presence of OH symptoms: asymptomatic (n=86) and symptomatic groups (n=86) (Table 1). No significant differences were observed between the two groups in terms of sex, age, BMI, underlying diseases (hypertension, diabetes, and coronary heart disease), clinical scale scores (MDS-UPDRS, MoCA, MMSE, and PDQ-39), or baseline hemodynamic parameters (SBP, DBP, PSV, EDV, and heart rate) (all P > 0.05). Compared to the asymptomatic group (0 points on the OH Symptom-Based Risk Score; Table 1), the symptomatic group showed significantly greater reductions in all parameters measuring the extent of BP and CBFv decline after standing (ΔSBPmax, ΔSBPmean, ΔDBPmax, ΔDBPmean, ΔPSVmax, ΔPSVmean, ΔEDVmax, ΔEDVmean) (all P < 0.001). Among the dCA parameters, VLF and LF gains were significantly higher in the symptomatic group than those in the asymptomatic group (P < 0.05), indicating impaired dCA function. Furthermore, the proportion of symptomatic patients with initial OH, nOH, and MSA was significantly higher than those with classical OH (64.6% vs. 31.9%, P < 0.001), non-nOH (57.5% vs. 37.9%, P = 0.012), and those with PD (61.8% vs. 42.3%, P = 0.013), respectively.

**Table 1.**
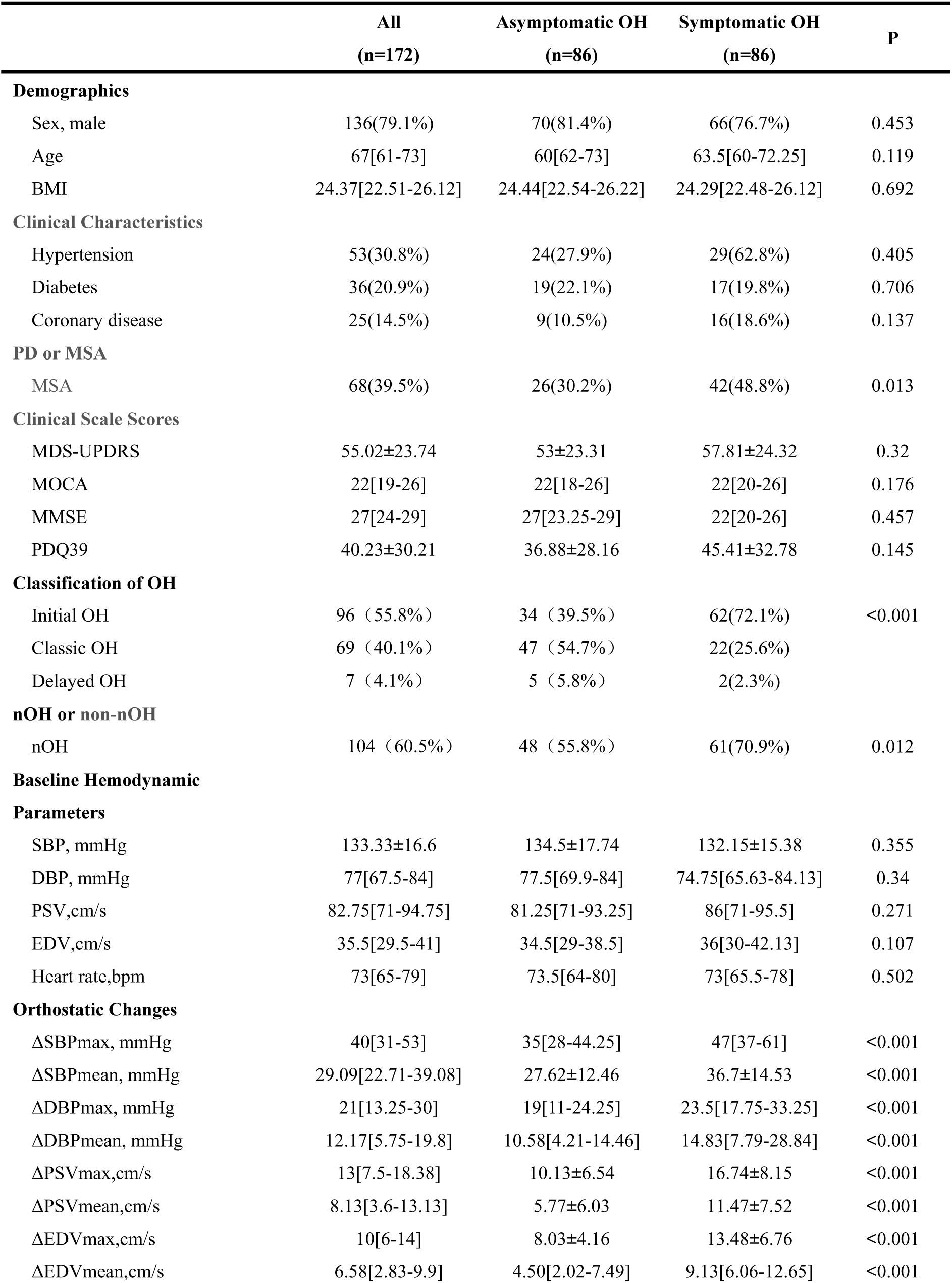
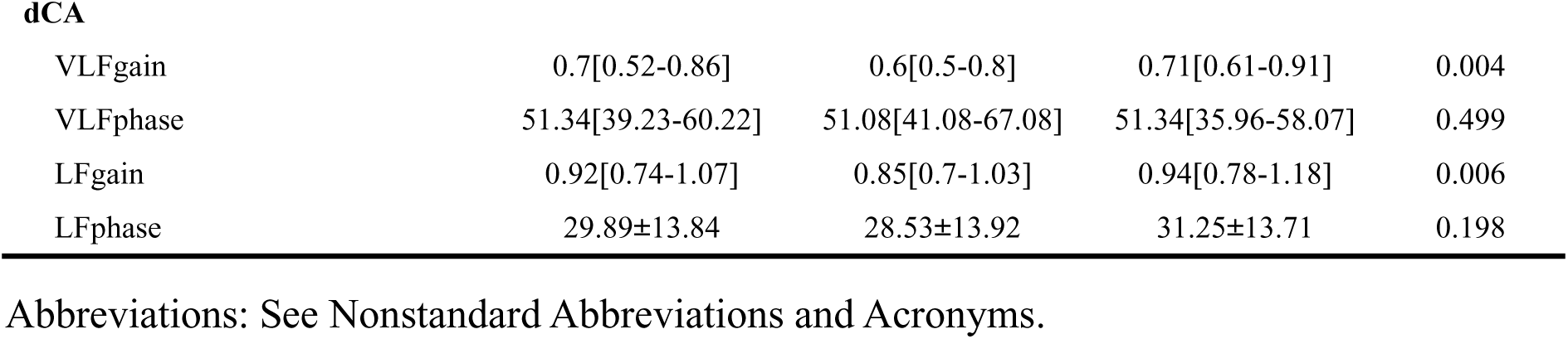
Demographic and clinical data in asymptomatic and symptomatic OH groups.

### Comparison of characteristics among patients with different OH Symptom-Based Risk Scores

Table 2 presents the characteristics of patients across different OH Symptom-Based Risk Score groups. Patients were stratified into three groups: 0 points (asymptomatic, n=86), 1 point (n=53), and 2 points (n=33). Significant differences were observed in the proportion of coronary heart disease between patients scoring 2 points and asymptomatic patients, and in MMSE scores between patients scoring 2 and 1 points (both P < 0.05). However, no significant differences were observed in sex, age, BMI, hypertension, diabetes, scores on other clinical scales, or baseline hemodynamic parameters (all P > 0.05). All parameters measuring the extent of BP and CBFv decline after standing showed progressively greater reductions with increasing OH Symptom-Based Risk Scores. VLF gain and LF gain were significantly lower in asymptomatic patients than in those with scores of 1 or 2 (all P < 0.05). Figure 2 displays proportions of the OH Symptom-Based Risk Scores across OH subtypes and synucleinopathy types, revealing significantly higher OH Symptom-Based Risk Scores in patients with initial OH (vs. classic OH), nOH (vs. non-nOH), and MSA (vs. PD) (all P < 0.05). Within the symptomatic OH cohort, we compared patients scoring 2 with those scoring 1 point to predict high-risk patients susceptible to critical events. All parameters measuring the extent of BP and CBFv decline after standing were significantly higher in patients scoring 2 points than in those scoring 1 point (all P < 0.05); however, no differences were observed in VLF or LF gain (both P > 0.05). The proportion of patients with a score of 2 was significantly higher in patients with initial OH and MSA than in those with classic OH (48.4% vs. 13.6%, P = 0.004) and PD (50% vs. 27.3%, P = 0.03). In contrast, patients with a score of 2 showed a higher prevalence of nOH than those with a score of 1(41.7% vs. 30.8%) without statistical significance (P = 0.34).

**Figure 2.**
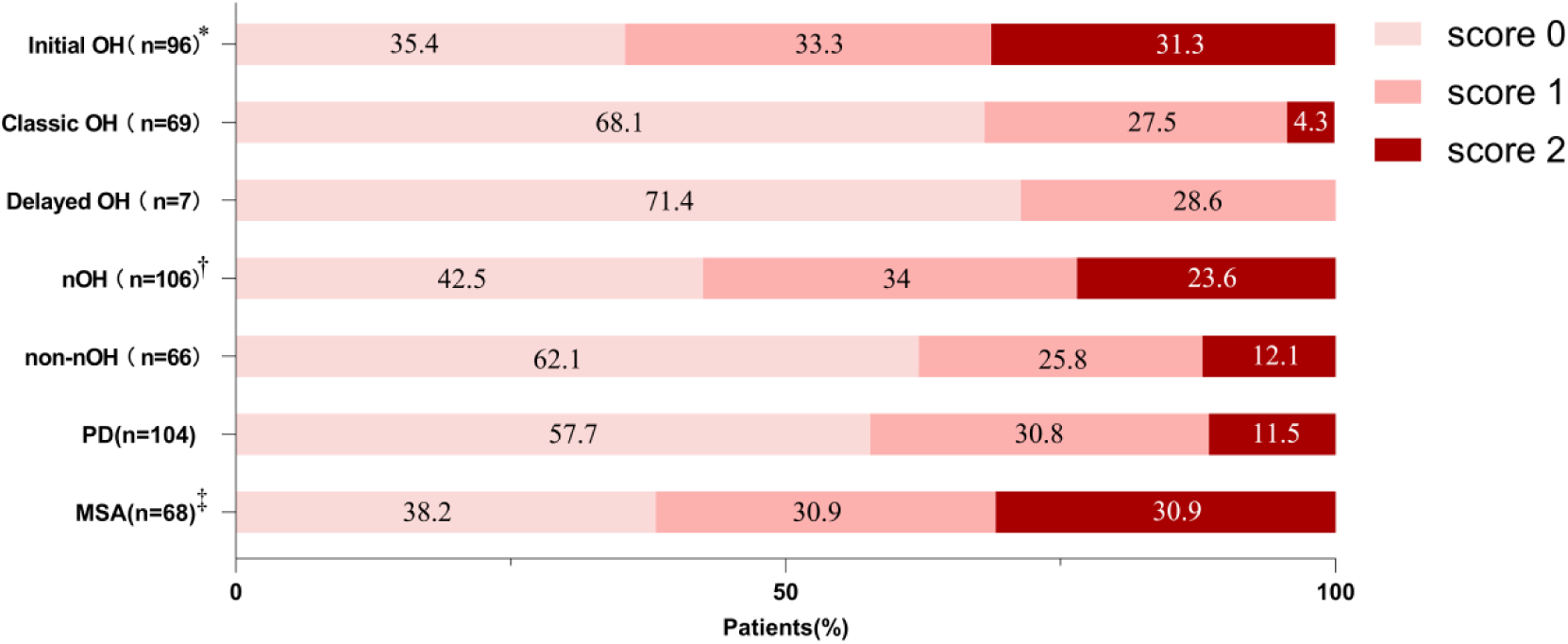
OH Symptom-Based risk scores by OH classification and α-synuclein type, the numbers on different bars represent percentages. nOH: Neurogenic orthostatic hypotension; PD: Parkinson’s disease; MSA: multiple system atrophy. * *P* <0.05 initial OH versus classic OH in OH symptom score. † *P* <0.05 nOH versus non-nOH in OH symptom score. ‡ *P* <0.05 MSA versus PD in OH symptom score.

**Table 2.** Comparisons of various factors between different OH Symptom-Based risk scores groups.

|  | OH Symptom-Based risk score |  |  | P |
| --- | --- | --- | --- | --- |
|  | 0 (n=86) | 1 (n=53) | 2 (n=33) |  |
| Demographics |  |  |  |  |
| Sex, male | 70(81.4%) | 39(73.6%) | 27(81.8%) | 0.498 |
| Age | 60[62-73] | 65[61-74] | 62[59.5-70.5] | 0.19 |
| BMI | 24.44[22.54-26.22] | 24.77[22.93-26.12] | 23.12[21.95-25.93] | 0.22 |
| Clinical Characteristics |  |  |  |  |
| Hypertension | 24(27.9%) | 16(30.2%) | 13(39.4%) | 0.551 |
| Diabetes | 19(22.1%) | 7(13.2%) | 10(30.3%) | 0.197 |
| Coronary disease | 9(10.5%) | 6(11.3%) | 10(30.3%)* | 0.025 |
| Clinical Scale Scores |  |  |  |  |
| MDS-UPDRS | 53±23.31 | 55.85±23.39 | 65±27.74 | 0.566 |
| MOCA | 22[18-26] | 23[20.25-26.75] | 21[19-25] | 0.097 |
| MMSE | 27[23.25-29] | 27[26-28] | 26[23-27]† | 0.174 |
| PDQ39 | 36.88±28.16 | 44.76±34.89 | 47.36±26.83 | 0.22 |
| Baseline Hemodynamic Parameters |  |  |  |  |
| SBP, mmHg | 134.5±17.74 | 132.32±14.78 | 131.88±16.55 | 0.772 |
| DBP, mmHg | 77.5[69.9-84] | 73.5[64.25-84.25] | 76.5[66.5-83.25] | 0.63 |
| PSV,cm/s | 81.25[71-93.25] | 90[69.75-97.25] | 82.5[75-92] | 0.494 |
| EDV,cm/s | 34.67±10.01 | 35.77±8.8 | 37.15±9.26 | 0.253 |
| Heart rate,bpm | 74.14±12.71 | 70.17±8.84 | 74.72±8.45 | 0.95 |
| Orthostatic Changes |  |  |  |  |
| ΔSBPmax, mmHg | 35[28-44.25] | 42[34-55]* | 53[43-66.5]*† | <0.001 |
| ΔSBPmean, mmHg | 27.62±12.46 | 31.94±13.04 | 44.34±13.66*† | <0.001 |
| ΔDBPmax, mmHg | 19[11-24.25] | 21[13.5-28.5] | 30[22.5-38]*† | <0.001 |
| ΔDBPmean, mmHg | 10.58[4.21-14.46] | 11.17[4.92-16.25] | 28.67[15.32-47.75]*† | <0.001 |
| ΔPSVmax,cm/s | 10.13±6.54 | 14.4±7.64* | 20.52±7.59*† | <0.001 |
| ΔPSVmean,cm/s | 5.77±6.03 | 8.98±6.68* | 15.47±7.14*† | <0.001 |
| ΔEDVmax,cm/s | 8.03±4.16 | 12.20±6.31* | 15.55±7.04*† | <0.001 |
| ΔEDVmean,cm/s | 4.50[2.02-7.49] | 7.67[5.42-10.58]* | 11.42[7.58-14.21]*† | <0.001 |
| dCA |  |  |  |  |
| VLFgain | 0.6[0.5-0.8] | 0.71[0.57-0.91]* | 0.71[0.64-0.95]* | 0.11 |
| VLFphase | 51.08[41.08-67.08] | 51.42[35.87-60.25] | 51.34[36.96-54.13] | 0.582 |
| LFgain | 0.85[0.7-1.03] | 0.97[0.76-1.18]* | 0.94[0.81-1.16]* | 0.025 |
| LFphase | 28.53±13.92 | 32.19±14.59 | 29.72±12.22 | 0.453 |

Abbreviations: See Nonstandard Abbreviations and Acronyms.
\* $P < 0.05$ versus patients with OH symptom score 0.
† $P < 0.05$ versus patients with OH symptom score 1.

### Multivariable models for OH symptom identification and risk stratification

During model development to identify symptomatic patients, the initial OH and dCA parameters were excluded because of high multicollinearity. Since model outcomes using ΔDBP instead of ΔSBP yielded comparable results (all P < 0.05), only the models with ΔSBP are presented. Four models were constructed: Model 1: adjusted for clinical data (sex, age, BMI, MSA or not, nOH or not), ΔSBPmax and ΔPSVmax; Model 2: adjusted for clinical data, ΔSBPmean and ΔPSVmean; Model 3: adjusted for clinical data, ΔSBPmax and ΔEDVmax; Model 4: adjusted for clinical data, ΔSBPmean and ΔEDVmean. The binary logistic regression results are presented in Table 3. The ROC curves and bootstrap validation results for each model are shown in Figure 3. Models using maximum decline values (Models 1 and 3) demonstrated superior performance compared with those using mean values (Models 2 and 4). Model 3 (ΔSBPmax and ΔEDVmax) demonstrated optimal performance with an AUC of 0.788 [95% CI: 0.72–0.856], sensitivity 64%, specificity 85%, bootstrap-validated accuracy 74.5% [68–80.8%], and F1 score 0.71.

**Figure 3.**
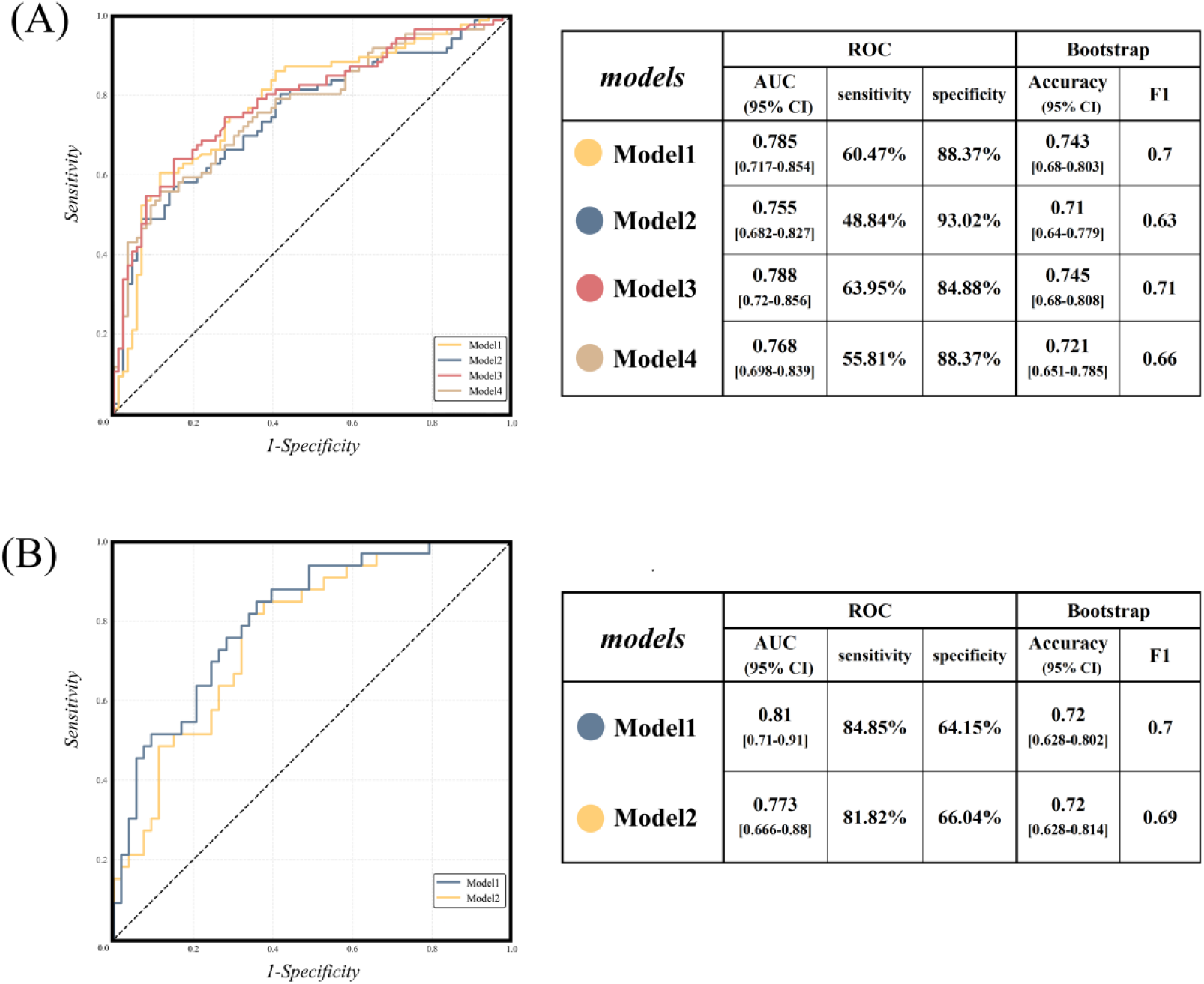
ROC curves and Bootstrap results for binary logistic regression models in (A) identifying asymptomatic and symptomatic OH patients;(B)stratifying symptomatic OH patients by critical symptom risk. Abbreviations: See Nonstandard Abbreviations and Acronyms. (A) Model1: Adjusted for clinical data (sex, age, BMI, MSA or not, nOH or not), ΔSBPmax and ΔPSVmax; Model2: Adjusted for clinical data, ΔSBPmean and ΔPSVmean; Model3: Adjusted for clinical data, ΔSBPmax and ΔEDVmax; Model4: Adjusted for clinical data, ΔSBPmean and ΔEDVmean. (B) Model1: Adjusted for clinical data (sex, age, BMI, MSA or not), ΔSBPmean and ΔPSVmean; Model2: Adjusted for clinical data, ΔSBPmean and ΔEDVmean.

**Table 3.** Binary logistic regression models in identifying asymptomatic and symptomatic OH patients.

|  |  | **OR (95% CI)** | **P** |
| --- | --- | --- | --- |
| Model1 | ΔSBPmax | 1.04 (1.02-1.07) | 0.001 |
| | ΔPSVmax | 1.12 (1.06-1.17) | <0.001 |
| Model2 | ΔSBPmean | 1.04 (1.01-1.07) | 0.007 |
| | ΔPSVmean | 1.11 (1.05-1.18) | <0.001 |
| Model3 | ΔSBPmax | 1.04 (1.01-1.07) | 0.002 |
| | ΔEDVmax | 1.18 (1.10-1.27) | <0.001 |
| Model4 | ΔSBPmean | 1.03 (1.01-1.06) | 0.02 |
| | ΔEDVmean | 1.20 (1.10-1.31) | <0.001 |

Abbreviations: See Nonstandard Abbreviations and Acronyms.
Model1: Adjusted for clinical data (sex, age, BMI, MSA or not, nOH or not), ΔSBPmax and ΔPSVmax; Model2: Adjusted for clinical data, ΔSBPmean and ΔPSVmean; Model3: Adjusted for clinical data, ΔSBPmax and ΔEDVmax; Model4: Adjusted for clinical data, ΔSBPmean and ΔEDVmean.

The binary logistic regression results to predict high-risk patients susceptible to critical events among symptomatic patients (OH Symptom-Based Risk Scores 1–2) are presented in Table 4. Only two models exceeded 70% bootstrap-validated accuracy: Model 1: adjusted for clinical data (sex, age, BMI, MSA or not), ΔSBPmean and ΔPSVmean; and Model 2: adjusted for ΔSBPmean and ΔEDVmean. The ROC curves and bootstrap results for both models are shown in Figure 3. Both models demonstrated comparable performance using the mean values. Model 1 (ΔSBPmean and ΔPSVmean) showed superior performance with an AUC of 0.81 [95% CI: 0.71–0.91], sensitivity 85%, specificity 64%, bootstrap-validated accuracy 72% [62.8–80.2%], and F1 score 0.7 compared to Model 2.

**Table 4.** Binary logistic regression models in stratifying symptomatic OH patients by critical symptom risk.

|  |  | **OR(95%CI)** | **P** |
| --- | --- | --- | --- |
| Model1 | ΔSBPmean | 1.06 (1.02-1.10) | 0.003 |
| | ΔPSVmean | 1.13 (1.04-1.22) | 0.003 |
| Model2 | ΔSBPmean | 1.06 (1.02-1.10) | 0.004 |
| | ΔEDVmean | 1.10 (1.00-1.21) | 0.053 |

Abbreviations: See Nonstandard Abbreviations and Acronyms.
Model1: Adjusted for clinical data (sex, age, BMI, MSA or not), ΔSBPmean and ΔPSVmean;
Model2: Adjusted for clinical data, ΔSBPmean and ΔEDVmean.

## Discussion

This study enrolled 217 patients with OH, representing the largest cohort to date investigating OH in a-synucleinopathies. Methodologically, previous OH studies predominantly employed cuff-based measurements and tilt-table testing, typically utilizing intermittent and short-duration BP monitoring, with rare simultaneous monitoring of CBFv changes in postural changes. In contrast, this study implemented continuous NIBP, CBFv, and heart rate monitoring throughout the procedure, collecting multi-time point BP and CBFv data, and quantifying dCA parameters. Furthermore, the model construction revealed the necessity of multiparametric analysis for OH research and highlighted the value of standardized OH measurement protocols and devices. From a design perspective, we pioneered a risk stratification framework that classifies OH symptoms into distinct severity levels, because effective OH management critically depends on symptom risk severity and frequency^27^. Consequently, precise symptom risk stratification is essential for maximizing patient benefits.

Large-scale studies focusing on the association between BP decline and OH symptom severity are scarce. Freeman et al.^9^ conducted tilt table testing in 89 patients using intermittent BP measurements at 1-min intervals. Specifically, they performed detailed and independent analyses of different OH symptoms based on patients’ subjective reports of light-headedness, dizziness, and impending blackout, and evaluated symptom scores separately. Their analysis found no correlation between the extent of the BP drop or the lowest BP within the first 3 min of tilting and the severity of any of these three OH symptoms. Notably, intermittent BP measurements at 1-min intervals can easily miss immediate BP changes after postural changes, especially in patients with initial OH and substantial BP drops. Furthermore, restricting symptom assessment to only the first 3 min post-tilting provides an inadequate observation window, potentially underestimating the initial/delayed OH prevalence and compromising symptom reproducibility. Accordingly, Palma et al.^28^ utilized intermittent electronic sphygmomanometer measurements at 1-min intervals to assess OH symptoms 3 min after postural change. They proposed an MAP < 75 mmHg at 3 min as a reliable predictor of OH symptoms. However, this fixed MAP threshold disregards the influence of circadian fluctuations and individual baseline BP variations. In contrast, this study utilized parameters reflecting BP decline magnitude post-postural change (ΔSBPmax and ΔSBPmean) through real-time beat-to-beat monitoring at multiple timepoints. Extending the monitoring duration to 10 min after standing enabled a more balanced characterization of the dynamic BP profiles.

Patients with OH often experience reduced cerebral perfusion due to abrupt decreases in blood pressure while standing. However, previous OH studies have largely overlooked the impact of cerebral hemodynamics (e.g., CBFv) in affected patients. A meta-analysis of three high-quality OH studies involving 177 patients showed that symptomatic patients with OH exhibited greater CBFv decline than asymptomatic patients^17^. The CBFv decline indirectly reflects cerebral hypoperfusion, which is mechanistically linked to neurovascular mechanisms underlying OH symptoms, including widespread neuronal suppression throughout the brain, dysfunction of oculomotor nuclei^29^, and other pathological alterations that clinically manifest as dizziness and blurred vision, whereas syncope is related to an insufficient metabolic reserve in the brain^30^. Thus, our findings highlight the necessity of CBFv measurements in OH research. In both predictive models (for symptom presence and risk severity stratification), the CBFv parameters demonstrated OR values comparable to those of the BP metrics (Tables 3 and 4). This underscores the incompleteness of the univariate BP analysis and emphasizes the need for comprehensive multiparametric OH monitoring devices. Additionally, while previous studies have rarely examined PSV and EDV separately, our findings indicated equivalent diagnostic values for PSV and EDV. Through NIBP and CBFv signal analyses, we quantified dCA parameters in patients with OH, revealing significantly worse regulatory function in symptomatic than in asymptomatic patients. In cases of dCA impairment, neurovascular units in regions such as the vestibular and visual cortices may sustain persistent hypoperfusion^9^. The resulting ischemic neurovascular dysfunction could contribute to OH symptoms, including dizziness and blurred vision. Our preliminary small-sample study in patients with PD also observed a trend of dCA impairment in symptomatic patients with OH^31^.

According to the 2021 US and European consensus on OH^18^, OH is classified into three subtypes based on the magnitude and timing of BP decline: initial, classic, and delayed OH; however, their prognostic implications remain controversial^3,19^. Gibbons et al.^7^ used intermittent BP measurements at 1-min intervals for subtyping and found a higher 10-year mortality in classic OH (64%) than in delayed OH (29%), suggesting a worse prognosis for classic OH. Initial OH has received the least research attention, potentially owing to the requirements for sensitive equipment and some theories categorizing it as transient physiological dysregulation^19^. Stephen et al.^16^ employed high-frequency BP measurements at 25-s intervals, revealing that initial OH increases short-term (dizziness, OR=1.49) and long-term risks (falls, HR=1.22; fractures, HR=1.16). In our study, the initial OH group showed higher OH Symptom-Based Risk Scores than the classic OH group (Figure 2), with substantially higher proportions of symptomatic and high-risk patients, underscoring the need for greater focus on initial OH. Mechanistically, nOH results from structural autonomic nervous system damage, often due to α-synuclein aggregation^3,8^, whereas non-nOH is related to reversible causes, such as medications or functional hypovolemia. Accordingly, Gibbons et al.^7^ observed that nOH was an independent mortality risk predictor, with non-nOH mortality comparable to that in the age-matched general population. MSA-related OH stems from degeneration of the brainstem and spinal cord sympathetic neurons^5^, manifesting early and progressing rapidly, whereas PD-related OH often occurs in advanced disease stages^1^, is associated with peripheral autonomic neuropathy, and shows lower symptom severity. Consistent with these mechanisms, in this study, patients with nOH (vs. non-nOH) and MSA (vs. PD) exhibited higher OH Symptom-Based Risk Scores and a higher prevalence of symptoms.

To the best of our knowledge, this is the first study to establish models that use multiparametric indicators to identify OH symptoms in patients. We constructed models using the maximum and mean values of the BP and CBFv decline after standing. As the maximum decline values showed significant differences between symptomatic and asymptomatic patients, Models 1 and 3 demonstrated superior performance. Among them, Model 3 showed the best performance with high specificity (85%) but relatively low sensitivity (64%). We propose that this performance characteristic likely stems not entirely from model limitations but rather reveals a special high-risk subgroup among asymptomatic patients characterized by substantial BP drops. Freeman et al. ^9^ reported approximately 40% patients with SBP drops > 60 mmHg were asymptomatic, while in our cohort, 69% (59/86) of asymptomatic patients had ΔSBPmax > 30 mmHg. Such substantial BP fluctuations cause multi-organ damage^3^, including a reduced glomerular filtration rate and increased cardiovascular risk, indicating a high actual injury risk despite the absence of symptoms. Potential reasons for the absence of symptoms include (1) habituation or degeneration of central pathways responsible for hypotension perception^9^ (symptoms such as dizziness exist, but patients cannot perceive them) and (2) altered dCA ranges in patients with neurodegenerative diseases^11^. Consequently, these actually high-risk “false-negative” patients may be experiencing occult cerebral hypoperfusion and multi-organ damage. The combination of high specificity and moderate sensitivity in Model 3 suggests that the risk stratification of asymptomatic patients to distinguish true low-risk individuals from high-risk subtypes may be a key future research direction for OH. Building on the successful identification of symptomatic patients, this study further developed a risk event prediction model for critical symptoms, such as falls and syncope, filling a research gap in the OH symptom domain. The predictive power of the maximum decline values weakened because both BP and CBFv declined substantially in symptomatic patients. Therefore, we used the mean decline values across multiple time points, which better reflected cerebral perfusion changes throughout the examination and amplified the characteristics of high-risk patients.

Recent data show that compared to symptoms like dizziness and blurred vision, patients with OH with falls or syncope have a 3-fold higher fracture risk^32^ and 40–64% increased all-cause mortality^3^. Consequently, the effective identification of patients with different OH symptom risks and implementation of stepwise pharmacological interventions based on severity are crucial for timely personalized management. Both models showed high sensitivity (84.9% and 81.9%, respectively), providing the first objective basis for maximizing the identification of potential high-risk patients and helping improve the targeting and risk stratification of current OH treatment strategies. However, the specificity (64.2% and 66%, respectively) required improvement. Future studies could optimize the model performance by incorporating multidimensional indicators such as neuroimaging biomarkers and refined autonomic function assessments.

This study has some limitations that require consideration. First, CBFv measurement via TCD assumes constant middle cerebral artery diameter to estimate cerebral blood flow. However, this assumption may be compromised during the AST because of sympathetically mediated smooth muscle contractions^33^ and transducer angle displacement. We minimized these effects by maintaining constant room temperature and humidity and securing the headframe to prevent transducer movement. Second, we did not correlate the findings with critical long-term outcomes, such as neurofunctional decline and cardiovascular event rates. Future multicenter cohorts studies are with larger sample sizes are required to study high-risk subgroups among asymptomatic patients and identify additional predictive indicators for continuous model optimization; the ability to predict patients prone to developing symptoms and those susceptible to critical events will enable timely, stepwise individualized treatment based on symptom severity. This approach can improve quality of life and prevent short- and long-term OH complications. Our goal was to establish a comprehensive OH risk stratification system that could provide reliable clinical risk assessment tools.

## Conclusion

This study established predictive models based on multiparametric indicators, primarily parameters reflecting the magnitude of postural BP and CBFv decline, which successfully predicted symptom occurrence and risk severity in patients with OH. This represents the first model to provide an objective quantitative tool for OH risk assessment in patients with PD and MSA, with significant implications for early clinical warning and optimization of current OH treatment strategies. Future studies should expand the sample size and continuously refine the OH diagnostic models to establish a comprehensive OH risk stratification system.

## Data Availability

The data that support the findings of this study are available from the corresponding author upon reasonable request.

## Acknowledgements

The authors thank all the staff that were involved in this study, as well as the patients and their families for their participation and cooperation.

## Disclosures

None.

### Nonstandard Abbreviations and Acronyms;

OH: orthostatic hypotension
nOH: neurogenic orthostatic hypotension;
PD: Parkinson’s disease;
MSA: multiple system atrophy;
BMI: body mass index;
MDS-UPDRS: Movement Disorder Society-unified Parkinson’s disease rating scale
PDQ39: 39-item Parkinson’s disease questionnaire
MMSE: mini-mental state examination
MoCA: montreal cognitive assessment
TFA: transfer function analysis;
dCA: dynamic cerebral autoregulation;
VLF: very low frequency;
LF: low frequency
BP: blood pressure;
SBP: systolic blood pressure;
DBP: diastolic blood pressure;
MAP: mean arterial pressure;
CBFv: cerebral blood flow velocity.
PSV: peak systolic velocity
EDV: end diastolic velocity
ΔSBPmax, ΔDBPmax, ΔPSVmax, ΔEDVmax: maximum decrease in SBP, DBP, PSV and EDV after standing, respectively.
ΔSBPmean, ΔDBPmean, ΔPSVmean, ΔEDVmean: mean decrease in SBP, DBP, PSV and EDV at all measured time points,15s, 30s, 1min, 3min, 5min, 7min, after standing, respectively.

